# Overall Outlook on Life and Cardiovascular Health Behaviors among Adolescent and Young Adult Females: A Cross-sectional Study

**DOI:** 10.64898/2026.11.03.26367227

**Authors:** Sophie H. Allende-Richter, Catherine Stamoulis, Ariana Reynier-Hernandez, Courtney A. Brown, Zoë E. Pollock, Hannah Chiert, Holly Gooding, Sarah de Ferranti

## Abstract

**Background:** Cardiovascular disease is the leading cause of death among women, with risk factors commonly emerging during adolescence. Emerging evidence indicates that psychological factors, such as optimism, affect cardiovascular health, yet this remains understudied among young women. To assess the association between overall outlook on life and preventive cardiovascular health behaviors among young women.

**Methods:** We conducted a secondary analysis of a cohort of 331 young women aged 15 to 24 years who completed the modified American Heart Association Women’s Health Survey (2017-2018), using multivariate regression adjusted for confounding.

**Results:** Over 50% of young women felt their outlook on life may impact their likelihood of developing a serious illness such as heart disease. Outlook on life was associated with prioritizing and taking care of health (β=0.18-0.19, 95% Confidence Interval (CI) [0.09, 0.29]; P=0.002) and usually following recommended healthy eating (β=0.12, 95% CI [0.03, 0.23]; P=0.016). Young women who identified as black (27.5%; N=91) felt that their outlook on life had a lower impact on their likelihood of developing a serious illness such as heart disease (Odds Ratio (OR)=0.80; 95% CI [0.65, 0.98]; P = 0.04), compared to other racial groups.

**Conclusion:** Our findings support previous observations indicating a cardioprotective role of optimism in cardiovascular health. However, this association may not hold among young women of color, potentially reflecting constraints in their lived environment that limit how much a positive outlook translates into perceived control over health outcomes. Strategies to reduce cardiovascular disease in women should address young women’s cardiovascular health and psychological and contextual factors, with particular attention to young women of color.

## Introduction

Cardiovascular disease is the leading cause of adverse pregnancy outcomes and deaths among women.^1,2^ Many young women are already at risk for such complications due to suboptimal cardiovascular health.^1,3^ The American Heart Association defines cardiovascular health based on eight metrics: four modifiable health behaviors (sleep, physical activity, smoking, and diet) and four health factors (body mass index (BMI), blood pressure, lipid levels, and serum glucose level). Ideal cardiovascular health is achieved when an individual or a population meets the recommended threshold on at least five metrics.^3,4^ Low cardiovascular health scores during childhood and adolescence predict poor cardiovascular health outcomes in adulthood.^4-6^ However, only 45% of US adolescents meet ideal cardiovascular health standards across five or more metrics, which drops dramatically to 20% among young women aged 18 to 39.^3,7^Our previously published work with a cohort of young women aged 15 to 24 revealed low cardiovascular disease awareness in this age group.^8,9^ Young women also cited stress and competing priorities as perceived barriers to adopting heart-healthy behaviors.^9^ These findings align with growing evidence linking psychological health factors to cardiovascular disease pathophysiology.^10^

For example, chronic stress exposure has been associated with decreased myocardial perfusion, arterial damage, and obesity.^11,12,13^ Psychological health factors may also operate bidirectionally; just as stress may harm cardiovascular health, a growing body of evidence indicates that positive psychological health attributes, such as optimism or outlook on life, are associated with fewer cardiovascular disease events and healthier cardiovascular trajectories in adult cohorts and may confer cardioprotective benefits.^10,14-16^ While adolescence and young adulthood are recognized as critical periods for cardiovascular health prevention, little is known about the protective psychological factors in this age group. Given that cardiovascular health trajectories are established early during puberty and that adult women bear a high burden of cardiovascular disease, including adverse pregnancy-related outcomes, identifying modifiable protective psychological factors during adolescence and young adulthood represents a critical opportunity in cardiovascular prevention strategies for women.^1,3,17^ Therefore, we sought to examine the relationship between young women’s overall outlook on life and their self-reported adherence to preventive cardiovascular health behaviors. We also examined this association in relation to socio-demographic factors.

## Methods

### Data source and study participants

We conducted a secondary analysis of a cohort of 331 young women who completed the modified American Heart Association (AHA) Women’s Health Survey for adolescent and young adult women (2017-2018).^8,18^ Study participants self-identified as cisgender women aged 15 to 24, spoke either English or Spanish, and received care at an urban academic hospital in Boston, Massachusetts, in its adolescent and preventive cardiology outpatient specialty clinics and at a community primary care clinic from September 2017 to January 2018.^8^ A bilingual or English-speaking research assistant approached eligible patients in the exam room or clinical area before their encounter with the provider and enrolled them in the primary study after obtaining informed consent. Participants completed the survey anonymously on an iPad.

### Measures and Study Design

The primary study survey instrument was adapted from the AHA National Women’s Health Study (Appendix Table 1) and pilot-tested with the Hospital Teen Advisory Board.^8,18 19^It encompasses the following domains: (1) cardiovascular disease awareness, (2) preventive health behaviors, (3) sources of heart health information, (4) barriers to adopting preventive health behaviors, and (5) access to preventive health behaviors. We also collected the following participant characteristics: gender, age, race (categorized as black, white, or other; including participants who identified as another race or more than one race), and Hispanic ethnicity. Socio-economic indicators included self-reported household income (an ordinal measure ranging from (<$15,000 (=1) to ≥$250,000 (=11)), health insurance coverage status as a binary variable, and caregiver education level (an ordinal measure from less than high school (=1) to completed graduate school (=8).^19^ Participants’ medical history included self-reported history of high cholesterol, prediabetes or diabetes mellitus, depression, smoking status, family history of heart disease, and body mass index (BMI). BMI was calculated from self-reported height and weight (kg/m^2^). Participants were classified as overweight if the BMI was ≥ 85th percentile and < 95th percentile for age (if under 20 years) or if the BMI was ≥ 25 and < 30 kg/m2 (if 20 years of age and older), and with obesity if the BMI was ≥ 95th percentile for age (if under 20 years) or if the BMI was ≥ 30 kg/m2 (if aged 20 years and older).^20,21^ We surveyed young women regarding their overall outlook on life and adherence to cardiovascular health recommendations. We assessed their responses to the question *“In general, would you say your overall outlook on life is”* using an ordinal scale from poor (=1) to excellent (=5). To determine the main factors influencing their outlook, we used a dichotomous (yes/no) response to the question *“Which of the following have the biggest impact on your overall outlook on life?”* with options including managing school, family, and work commitments; physical, emotional, and family health; financial stability concerns; time to complete necessary tasks; caring for children and/or an adult family member; and spiritual life. We evaluated their perceptions of how their overall outlook on life impacts their health in response to the question: *“How much do you think overall outlook on life impacts your likelihood (1) to develop a serious illness like heart disease; (2) to successfully manage a serious illness like heart disease if you develop it”*, using a Likert scale. Finally, we assessed their agreement with various statements about cardiovascular health recommendations: “*Please indicate how much you agree or disagree with the following statements: I don’t get enough sleep regularly; I am taking care of my health; my health is a priority; I am so busy taking care of others; I don’t take good care of myself; I usually follow healthy eating habits; when life gets busy, I skip exercise; my muscles and joints ache regularly; I am concerned about my alcohol intake*.*”*

### Statistical Analysis

Depending on the outcome, we developed multivariate generalized linear or logistic regression models to examine relationships among participants’ overall outlook on life, self-reported adherence to preventive cardiovascular health behaviors, and socioeconomic factors. We adjusted the models for potential confounding by age, race, ethnicity, BMI, and depression. P-values for multiple cardiovascular health questions were adjusted for the False Discovery Rate using the Benjamini-Hochberg approach.^22^

### Ethics

The Hospital Institutional Review Board approved this study.

## Results

331 participants completed the survey, and their characteristics are summarized in Table 1

**Table 1:** Survey respondents’ characteristics. *All personal medical history variables were self-reported. Classification of overweight/obesity in the AYA sample was based on pediatric (if under age 20 years) and adult (if 20 years of age and older) Centers for Disease Control body mass index thresholds using self-reported height and weight for the AYA sample. Classification of overweight/obesity in the AHA sample was based on self-report of being 20lb or more over one’s ideal weight for height. Personal history of heart disease was considered present if the participant noted any history of a heart attack or stroke*.

| **Participants Characteristics** | **Number (%)** |
|---|---|
| Age |  |
| 15-17y | 91 (27.5) |
| 18-21y | 140 (42.3) |
| 22-24y | 69 (20.8) |
| Missing | 31 (9.4) |
| Race/Ethnicity |  |
| Black | 91 (27.5) |
| White | 74(22.4) |
| Other (including mixed race) | 112 (33.8) |
| Hispanic | 123 (37.2) |
| Missing race or ethnicity | 47 (14.3) |
| Household income |  |
| \$< 35,000 | 105 (31.7) |
| 35,000 - <50,000 | 14(4.2) |
| 50,000 – <75,000 | 17 (5.1) |
| ≥75,000 | 38 (11.5) |
| Don't know/Missing | 157 (47.5) |
| Caregiver education |  |
| Less than high school | 21 (6.3) |
| Completed some high school | 29 (8.8) |
| Completed high school | 49 (14.8) |
| Completed some college | 29 (8.8) |
| Associates Degree | 30 (9.1) |
| Completed College | 54 (16.3) |
| Completed some graduate school | 8 (2.4) |
| Completed graduate school | 69 (20.8) |
| Unsure/Missing | 42 (12.7) |
| Health insurance status |  |
| Yes | 301 (90.9) |
| No | 1 (0.3) |
| Missing | 29 (8.8) |
| Personal medical history |  |
| Overweight/obesity | 85 (25.7) |
| Depression | 107 (32.3) |
| Smoking | 23 (7.0) |
| Family History of Heart disease | 62 (18.7) |
| High blood pressure | 23 (9.0) |
| High cholesterol | 17 (5.1) |
| Diabetes/pre-diabetes | 17 (5.1)/22 (6.7) |
| Missing | 52 (15.7) |

### Overall Outlook on life and influential factors

Young women rated their overall outlook on life as *excellent* (8%, N=26), *very good* (27%, N=82), *good* (35%, N=107), *fair* (26%, N=81), or *poor* (4%, N=11). The distribution of these findings is shown in Figure 1. Young women identified their emotional health (53%, N=175), their ability to manage school commitments (52%, N=172), and their physical health (41%, N=136) as the top three most influential factors that had *the biggest impact on their overall outlook on life*, as shown in Figure 2. Participant education level was collinear with age; thus, associations with these parameters were tested in separate models. Young women’s overall outlook on life was positively associated with participant age and education level (β = 0.06, 95% CI = [0.01, 0.11], p = 0.03 for age, and β = 0.12, 95%CI = [0.02, 0.21], p = 0.01 for education level), and negatively associated with depression (β = -0.61, 95% CI = [-0.86, -0.36, p<0.01). Associations with race, ethnicity, or BMI were nonsignificant (p>0.1). Finally, overall outlook on life was also positively associated with caregiver education level (β = 0.08, 95% CI = [0.03, 0.14], p<0.01).

**Figure 1.**
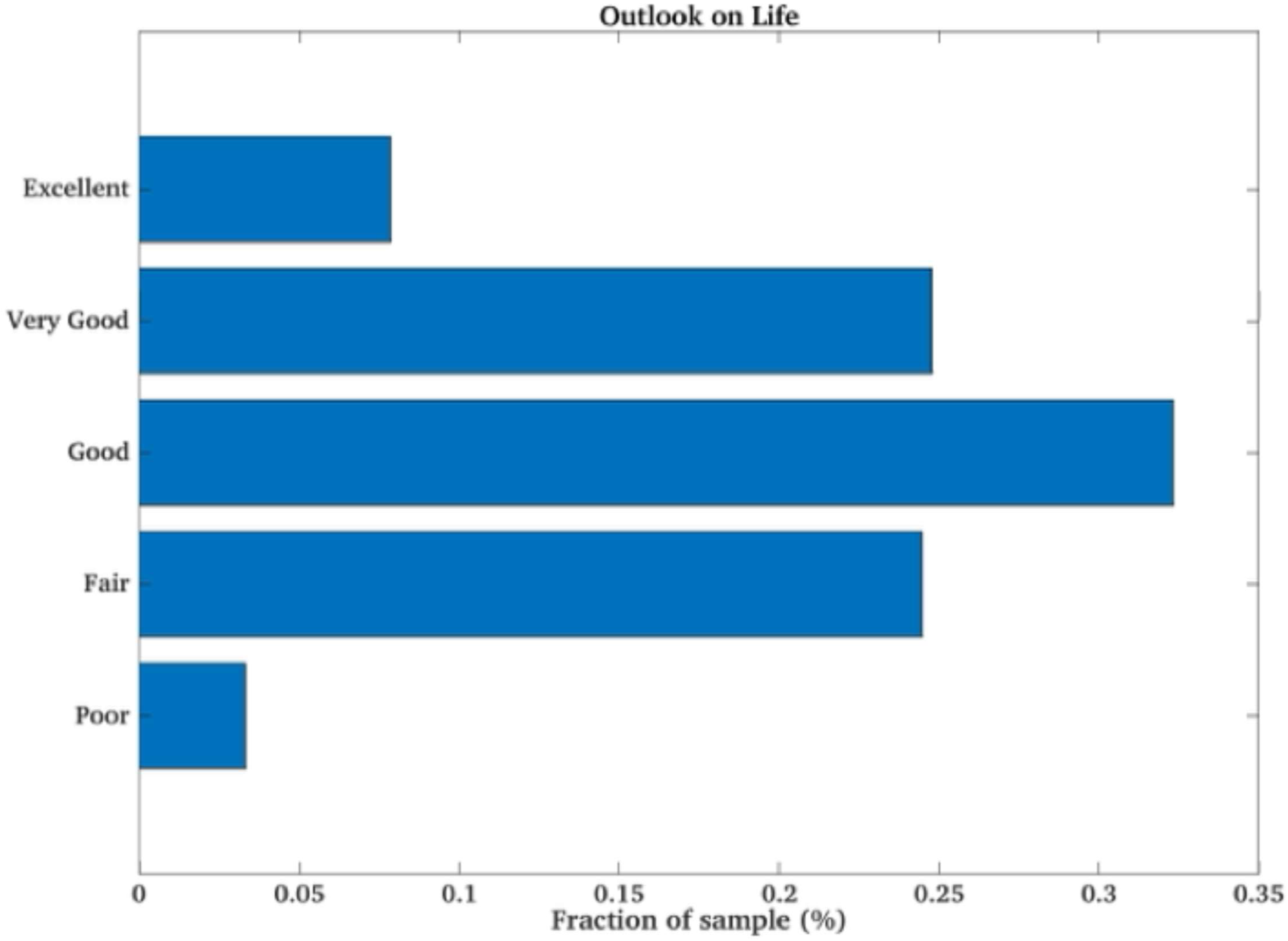
Self-Rating of Young Women’s Overall Outlook on Life

**Figure 2.**
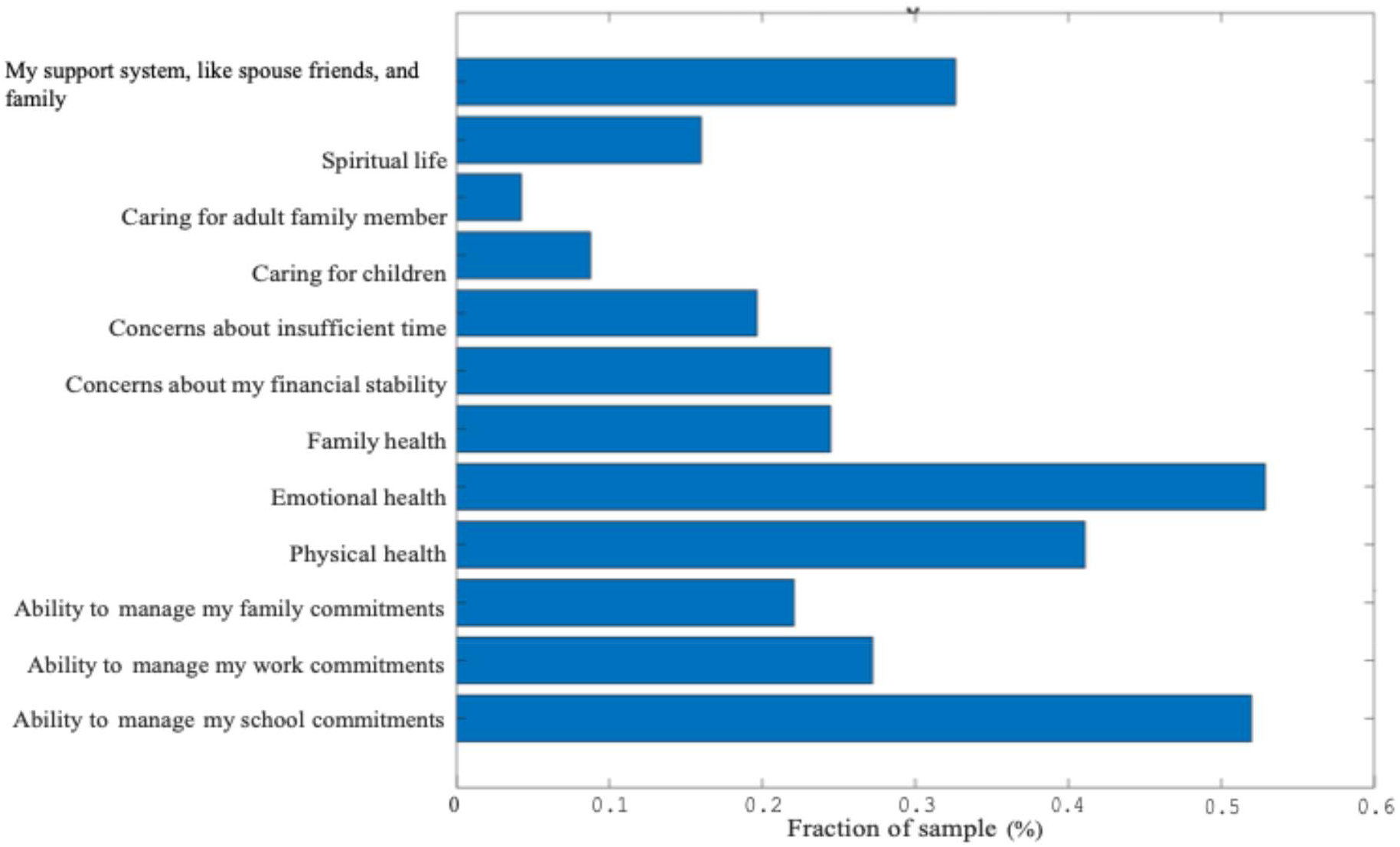
Young Women’s Perceived Influential Factors on Outlook on Life

### Young women’s perception of overall outlook impact on health outcomes

Approximately 55% of young women (N=295) affirmed that their *overall outlook on life had an impact on their likelihood of developing a serious illness like heart disease*, with responses ranging from *“a great deal”* (4%, N=11), *“very much”* (9%, N=27), to *“some extent”* (42%, N=123). By contrast, 45% (N=134) reported that their *overall outlook on life did “not at all” have an impact on their likelihood of developing a serious illness like heart disease*. Similarly, 69% of young women (N=294) believed that *their overall outlook on life had an impact on their likelihood of successfully managing a serious illness like heart disease*; responses ranged from “*a great deal*” (13%, N=38), “*very much”* (21%, N=62), to “*some extent*” (35%, N=102).

In this analysis, compared to women under the age of 18 (27%; N=91), young women aged 18 years and older (63%; N=209) were more likely to report a higher impact of outlook on life on their likelihood of developing (OR=1.45; 95% CI [1.17, 1.80]; P<0.01). Age differences in the impact of outlook on life on managing serious illness, such as heart disease, were nonsignificant (P = 0.09). Young women who identified as black (27.5%; N=91) reported that their *overall outlook on life had lower impact on their likelihood of developing a serious illness* (OR=0.80; 95% CI [0.65, 0.98]; P = 0.04) compared to other racial groups. Differences between racial groups in the impact of outlook on life on *successfully managing a serious illness like heart disease* were nonsignificant (P = 0.60). Similarly, there were no statistical differences between Hispanic and non-Hispanic young women on the impact of outlook on life on developing or managing serious illness (P>0.20) compared to other races.

### Outlook on life and self-reported statements of cardiovascular health behaviors and physical health

Young women were surveyed on their adherence to cardiovascular health behaviors and measures of physical health. Regarding cardiovascular health behaviors, 87% (N=252) strongly or somewhat agreed that their *health is a priority*, and 62.5% (N=185) strongly or somewhat agreed *that they usually follow recommended healthy eating habits*. While 68% (N=203) acknowledged *not getting enough sleep regularly*, 73% (N=216) reported that *exercise was the first activity they skip when life gets busy*, and 38% (N=112) *reported experiencing muscle and joint aches regularly*.

Prior to examining associations with overall outlook on life, we assessed whether these statements varied by socioeconomic and racial/ethnic factors. Young women’s statement of *following recommended healthy eating habits* was positively associated with caregiver education level (P<0.01). Compared with white participants, young women of other races and those who self-identified as black were more *likely to report being so busy taking care of everyone else instead of themselves* (P<0.01), *not getting enough sleep* (P=0.01), *experiencing regular muscle and joint aches* (P=0.02), and *being concerned about their alcohol intake* (P=0.04).

#### Outlook on life and statement of cardiovascular health

We then examined the association between young women’s overall outlook on life and their self-reported statement of cardiovascular health behaviors and physical health, after adjusting for age, caregiver education level, BMI, and depression. Young women’s overall outlook on life was positively associated with their statements about their ability to prioritize their health (β=0.19, 95% CI [0.08, 0.29]; P<0.01) and to follow healthy eating recommendations (β =0.12, 95% CI [0.03, 0.23]; P =0.02). By contrast, young women’s overall outlook on life was negatively associated with statements about *not getting enough sleep on a regular basis* (β = -0.21, 95% CI [-0.35, -0.08]; P=0.02), *being so busy taking care of everyone else that I don’t take good care of myself* (β = -0.12, 95% CI [-0.24, -0.02]; P<0.01), and *my muscles and joints ache on a regular basis* (β = -0.16, 95% CI [-0.28, -0.04]; P = 0.01) (**Table 2**).

**Table 2.** Associations between outlook on life (the primary predictor) and statements related to cardiovascular health (adjusted for age, caregiver education, depression, and BMI) *Adjusted for the False Discovery Rate Outlook on life is on a scale of poor (=1) to excellent (=5). Responses are from strongly disagree (=1) to strongly agree (=4)

|  | **Regression Coeff (β) for Outlook on life** | **95%Confidence Interval (CI)** | **Adjusted P-value<sup>\*</sup>** |
|---|---|---|---|
| I don't get enough sleep on a regular basis | -0.21 | [-0.35, -0.08] | 0.016 |
| I am taking care of my health | 0.18 | [0.09, 0.28] | 0.002 |
| My health is a priority for me | 0.19 | [0.08, 0.29] | 0.002 |
| I’m so busy taking care of everyone else, I don’t take good care of myself | -0.12 | [-0.24, -0.02] | 0.002 |
| I usually follow recommended healthy eating habits | 0.12 | [0.03, 0.23] | 0.016 |
| My muscles and joints ache on a regular basis | -0.16 | [-0.28, -0.04] | 0.014 |

## Discussion

In this study, we examined a cohort of women aged 15 to 24 who completed the modified AHA National Women’s Health Survey (2017-2018) and received care in a pediatric and adolescent academic center in Massachusetts.^8^ We explored young women’s perceptions of their overall outlook on life and how these relate to their adherence to preventive cardiovascular health behaviors, as well as how socio-demographic indicators shape this relationship.

Our main findings indicate that most young women who completed this study self-rate their outlook on life positively. Young women identified their emotional and physical health, as well as their ability to manage their school commitments, as the top 3 contributors to their overall outlook on life. When examining socio-demographic correlates, overall outlook on life was associated with parental education level but not with race or ethnicity. More than 50% felt that their outlook on life had an impact on their likelihood of developing or managing a serious illness such as heart disease. Black and Hispanic respondents shared a different perception than white participants, stating that their outlook on life did not have any impact on their likelihood of developing or managing a serious illness.

Young women’s self-reported statement of cardiovascular health behaviors, in relation to their overall outlook on life self-rating, appeared to be more closely influenced by socio-economic indicators and contextual factors in their lived environment. Young women of other races and those who self-identified as Black were also more likely to report being so busy taking care of everyone else that they neglected their own health, not getting enough sleep, and experiencing poor physical health due to muscle and joint aches and thus difficulty adhering to recommended health behaviors. After adjusting for socio-economic indicators and race and ethnicity, young women’s overall outlook on life remained independently and positively associated with preventive cardiovascular health behaviors, as shown in our multivariate analysis. Overall outlook was positively associated with statements prioritizing health and eating well, and negatively associated with poor sleep, competing priorities in the lived environment, and poor physical health.

Taken together, our findings suggest that overall outlook on life is associated with preventive cardiovascular behaviors independent of socio-economic status, race, and ethnicity in young women aged 15 to 24. Our observations align with prior findings indicating an association between optimism and cardiovascular health trajectory in adult studies, suggesting a cardioprotective role of optimism detectable at an earlier age than previously described. ^10,14-16^ While this association was slightly attenuated by socio-economic indicators, the relationship in this study appears mediated by discrete factors in the lived environment that were more proximally influential, likely reflecting our participants’ developmental stage.^16^ This mediating effect may be especially pronounced among young women of color, possibly reflecting contextual constraints that limit the extent to which positive outlook translates into perceived control over health outcomes.

Despite these constraints, these findings indicate that young women perceive overall outlook on life as a relevant and potentially modifiable factor that could shape their health behaviors and engagement in care for cardiovascular disease prevention, trajectory, and management. This is particularly significant given the growing consensus that adolescence and young adulthood are key periods for cardiovascular prevention strategies.^1^ Understanding how psychological factors affect cardiovascular health in young women could have significant clinical implications for adult women’s cardiovascular prevention strategies, including screening practices and treatment response, by accounting for both psychological and contextual factors to support long-term cardiovascular risk reduction.

### Limitations

While these findings are encouraging and suggest new avenues in cardiovascular prevention strategies, we acknowledge several limitations in our study. A primary limitation is the cross-sectional design, which makes our findings observational and prevents establishing a causal relationship between overall outlook on life and cardiovascular health behaviors. Although caregiver education level is widely used to assess the relationship between socioeconomic circumstances and cardiovascular disease outcomes, our reliance on it as a proxy for socioeconomic status limited our ability to fully characterize socioeconomic influences on this relationship.^19^ Additionally, our reliance on self-reported data introduces potential recall and social desirability biases, particularly for sensitive items such as adherence to healthy eating or sleep habits. Furthermore, although our sample was adequately powered to detect the observed associations, it may not fully represent the broader population of young women, limiting generalizability. Lastly, our data were collected from a 2017-2018 AHA Women’s Health Survey dataset for young women; given evolving mental health trends among adolescents in subsequent years, including during the COVID-19 pandemic, more contemporary data may better capture current patterns in outlook on life and cardiovascular health behaviors.

In conclusion, our study findings indicate that overall outlook on life is associated with preventive cardiovascular health behaviors in young women and may confer cardioprotective benefits, with potential clinical implications for cardiovascular health prevention strategies. Future research should adopt a longitudinal study design with an adequately powered, nationally representative sample, incorporating more granular socioeconomic measures such as the Child Opportunity Index, along with depression and anxiety indices, as these conditions are prevalent among adolescents and potential confounders.^23^ This design would provide stronger data to assess long-term cardioprotective benefits and cardiovascular health trajectories associated with overall outlook on life among young women.

## Data Availability

A complete dataset can be provided upon request

## Acknowledgment

Funding supports: Dr. Allende-Richter was supported in part for this research by the P30HS029755 grant, funded by the Agency for Health Research and Quality (AHRQ) and the Patient-Centered Outcomes Research Institute (PCORI®), and by the COURAGE Program, funded by the National Institute of Diabetes and Digestive and Kidney Diseases (NIDDK) under award number U24DK132733.

Dr. Holly Gooding was supported for this research by K23HL122361, awarded by the National Institute of Lung and Blood.

## Abbreviations

AHA: American Heart Association
BMI: Body Mass Index
CI: Confidence Interval
OR: Odds Ratio

